# Knowledge, attitudes, and practices regarding priority zoonotic diseases (Mpox, Ebola, and Rabies) and associated factors among households in Bukavu, Democratic Republic of the Congo, 2025

**DOI:** 10.64898/2026.10.26.26366379

**Authors:** Arnold Kwibuka Murangaza, Chiribagula Zalinga Christian, Christian Mugisho Byamungu, Hugues Cakirwa Mandevu, Gentil Mwema Kisangani, Rosine Bigirinama Nshobole

## Abstract

**Background:** Zoonotic diseases are a major public health concern in the Democratic Republic of the Congo (DRC), where diseases such as rabies, Ebola, and Mpox pose ongoing health risks. Household knowledge, attitudes, and practices (KAP) play an important role in preventing zoonotic disease transmission. This study assessed household knowledge, attitudes, and practices regarding priority zoonotic diseases in Bukavu, South Kivu Province, DRC, and identified factors associated with satisfactory overall KAP.

**Methods:** A cross-sectional study was conducted from September 1 to 15, 2025, in the Bagira and Kadutu health zones of Bukavu. A total of 634 households were selected using two-stage cluster sampling. Data were collected using a structured questionnaire administered through KoboCollect and analyzed using Stata version 17. The questionnaire assessed knowledge, attitudes, and practices concerning Ebola, Mpox, and rabies. Knowledge was measured using 17 items, attitudes using five-point Likert-scale items, and practices using five behavioral items. Scores of at least 70% were classified as satisfactory. Descriptive statistics, group comparisons, and bivariate and multivariable logistic regression analyses were used. Nonparametric tests were used when appropriate. Associations were reported as odds ratios (ORs) with 95% confidence intervals (CIs), with p < 0.05 considered statistically significant.

**Results:** Overall, 42.9% of households demonstrated satisfactory knowledge of zoonotic diseases, 63.6% had favorable attitudes, and 11.7% demonstrated adequate preventive practices. Mpox was recognized as a zoonosis by 78.2% of respondents, Ebola by 66.6%, and rabies by 39.1%. Knowledge scores were higher among livestock-farming than non-livestock-farming households (p = 0.002). In the multivariable analysis of overall KAP, university-level education (OR = 12.06; 95% CI: 1.54–94.18; p = 0.018) and access to drinking water (OR = 6.74; 95% CI: 3.91–11.60; p < 0.001) were positively associated with satisfactory overall KAP.

**Conclusions:** Households in Bukavu reported generally favorable attitudes toward zoonotic disease prevention, but satisfactory knowledge was less widespread and adequate preventive practices were uncommon. Interventions should help translate awareness and favorable attitudes into practical preventive behaviors through targeted risk communication and health education, alongside improvements in access to drinking water and sanitation. A coordinated One Health approach involving human, animal, and environmental health stakeholders may strengthen household-level prevention of priority zoonotic diseases. The findings can help inform locally tailored prevention strategies and identify priorities for future community-based research.

## Background

Zoonotic diseases represent a significant global public health challenge, affecting billions annually and causing substantial human and economic losses(1,2) . They comprise infectious conditions that naturally transmit between vertebrate animals and humans, including bacterial diseases (brucellosis, anthrax, salmonellosis) and viral diseases (rabies, Ebola, Mpox, avian influenza) (2,3).

Zoonotic diseases are transmitted in various ways. Direct contact with the bodily fluids of infected animals (saliva, blood) is the main route of transmission, accounting for around 60% of cases. Vector transmission, involving mosquitoes and ticks in particular, is estimated at 17%. Consumption of contaminated food, such as raw milk or undercooked meat, accounts for nearly 20% of infections. Indirect transmission via contaminated environments or objects, on the other hand, remains less frequent, accounting for around 3% of cases.(3,4)

Each year, zoonoses affect nearly one billion people and cause millions of deaths, disproportionately impacting vulnerable populations in resource-limited regions (5,6). In response, the One Health approach emerged in 2005, offering an integrated vision of human, animal, and environmental health (4,7).

Africa and Asia are most affected due to high biodiversity, frequent human-animal contact, rapid urbanization, and limited surveillance resources(6,8) . In Africa, zoonoses represent approximately 32% of reported infectious disease outbreaks between 2001 and 2022, with a 63% increase over the past two decades(8,9).

Studies conducted in developing countries indicate low awareness of zoonoses despite their high prevalence. This situation is mainly associated with limited access to information, low literacy levels, and the priority given to other public health issues. Furthermore, the lack of effective communication strategies promotes the spread of these diseases and hinders the adoption of preventive measures(10–12).

In the Democratic Republic of Congo (DRC), zoonoses constitute a serious public health problem with frequent outbreaks of Ebola, Mpox, rabies, and avian influenza(13). The country has experienced 15 Ebola outbreaks since 1976 (14). In 2024, the Mpox epidemic tripled compared to 2023 (from 1,056 to 3,190 suspected cases), with case fatality increasing from 4.8% to 7.8%(13,15). Rabies remains endemic with mortality approaching 100% without post-exposure prophylaxis (16).

South Kivu Province, with its high population density and importance of livestock farming, is particularly exposed. The proximity between humans, domestic animals, and wildlife increases transmission risks, compounded by poverty and insufficient health services(17). Understanding households knowledge, attitudes, and practices (KAP) is essential to identify gaps and design appropriate interventions.

This study aimed to: (1) describe households’ knowledge, attitudes, and practices (KAP)levels regarding priority zoonoses; (2) compare KAP between livestock and non-livestock households; and (3) identify determinants of KAP levels in Bukavu city.

## Methods

### Study area

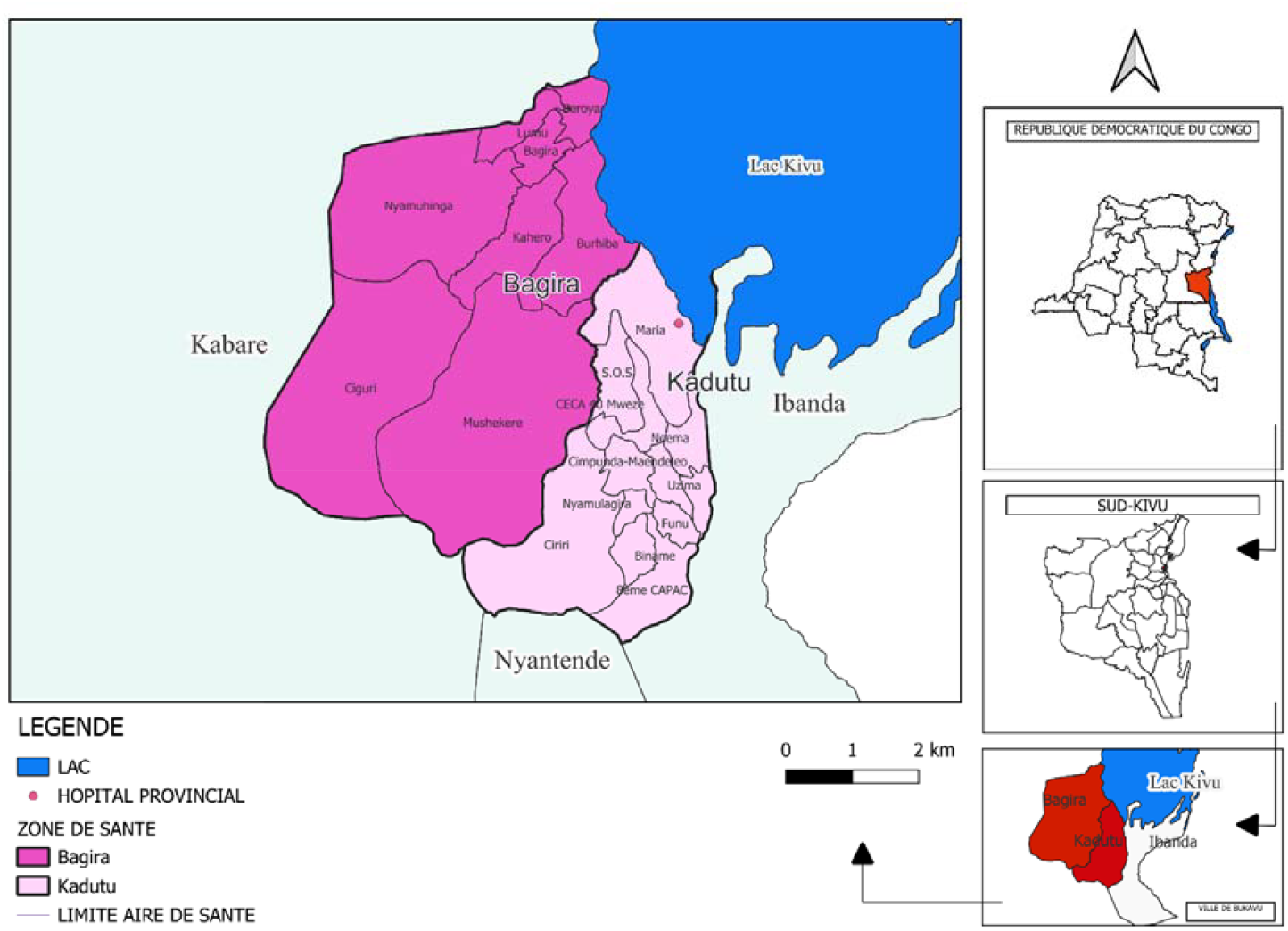

#### Study Design and Setting

A cross-sectional study was conducted from September 1–15, 2025, in Bukavu, the capital of South Kivu Province in eastern Democratic Republic of the Congo (DRC). The city has approximately 2,064,000 inhabitants with an estimated density of 19,833 inhabitants/km^2^ (18,19). South Kivu experiences a tropical highland climate and a complex ecosystem characterized by rich biodiversity, wildlife reservoirs such as rodents, bats, and primates, and close human-animal-environment interfaces exacerbated by ongoing deforestation and proximity to protected forest areas(20,21) . The region is also shaped by its aquatic environment notably Lake Kivu and urban river networks which heavily influences local trade, household water access, and sanitation dynamics(22).

The province faces persistent humanitarian crises, armed conflicts, and massive internal displacements, which weaken public health infrastructure and accelerate the re-emergence of zoonotic diseases(23–25), Beyond historical endemics such as rabies, brucellosis, and anthrax, South Kivu has recently been the epicenter of major outbreaks, including the 2023-2024 clade Ib mpox epidemic. which highlights the importance of strengthening community-level knowledge and preventive practices regarding zoonotic diseases.(26,27) , in a general context of low human development (28).

Within Bukavu, two contrasting health zones were selected to capture intra-urban socio-ecological heterogeneity: Kadutu, a highly congested urban health zone marked by dense informal markets, high human turnover, and active livestock trade(29) and Bagira, a peri-urban health zone characterized by lower population density, extensive household-level agropastoral activities (specifically pig and poultry farming), and direct spatial overlap with natural/rural ecosystems(30–33).

#### Study Population and Sampling

The target population comprised all households in Bagira and Kadutu health zones. Sample size was determined using Schwartz’s formula with design effect (DEFF = 1.5), yielding a minimum of 576 households. A 10% increase was applied for non-response, resulting in 634 households(34–36). Two-stage cluster sampling was used. In the first stage, 30 clusters were selected with probability proportional to size (PPS) in the health areas of Bagira (≈27%, 8 clusters) and Kadutu (≈73%, 22 clusters) using ENA-SMART software. At the second stage, households were selected by systematic sampling (37)from the household of the avenue chief. A sampling interval (k) was calculated based on the total number of households in each selected avenue, with the first household chosen randomly between 1 and k.

#### Data Collection

Data Collection were collected using a structured questionnaire administered via KoboCollect on Android tablets from 1st September to 15th September 2025 by trained data collectors under the supervision of the research team. The questionnaire was developed based on WHO KAP survey guidelines and validated tools from previous studies on zoonotic diseases (22,36–40). It covered sociodemographic characteristics (age, gender, marital status, education level, occupation, household size, and other relevant variables) and KAP regarding priority zoonoses (Ebola, Mpox, rabies) divided into three sections:

- Knowledge section assessing respondents’ knowledge of zoonotic diseases, including definitions, common examples, modes of transmission, risk factors, symptoms, and preventive measures;
- Attitude section evaluating perceptions regarding the seriousness of zoonotic diseases, perceived susceptibility, health-seeking behavior, and community prevention strategies;
- Practice section examining preventive behaviors and risk-related practices, including hygiene measures, animal handling practices, food preparation habits, and healthcare-seeking behavior following suspected exposure.

Knowledge was assessed through 17 items, attitudes via 5-point Likert scale items, and practices through 5 behavioral items. Scores were categorized as: <50% (poor), 50-69% (moderate), and ≥70% (satisfactory) (39–41).

Data were analyzed using Stata 17 after checking the completeness and consistency of the variables and recoding them. Knowledge, attitude, and practice (KAP) scores were calculated by assigning one point for each correct answer and then converting them into percentages. A threshold of 70% was used to classify participants as adequate or inadequate. The internal consistency of the items comprising the scores was assessed using Cronbach’s alpha coefficient.

Qualitative variables are presented as frequencies and percentages, while quantitative variables are described by mean ± standard deviation or median and interquartile range depending on their distribution, as assessed by the Shapiro -Wilk test. Comparisons between groups were performed using Student’s t-test or the Mann-Whitney/Kruskal-Wallis test, as appropriate.

The factors associated with CAP scores were explored using bivariate and then multivariate logistic regression. The results are reported as odds ratios (OR) with their 95% confidence intervals. The quality of model fit was assessed using the Hosmer–Lemeshow test, the area under the ROC curve, and the Link test. The threshold for statistical significance was set at p < 0.05.

#### Ethical Considerations

The study protocol was reviewed and approved by the Institutional Ethics Committee of the Catholic University of Bukavu (Comité d’Éthique de l’Université Catholique de Bukavu). The study was conducted in accordance with the Declaration of Helsinki. Verbal informed consent was obtained from all adult participants before data collection. Confidentiality and anonymity were maintained throughout the study.

### Results

#### Sociodemographic Characteristics

Of 634 participants, 50.5% were women and 58.0% were aged 18-35 years. Secondary education predominated (50.5%). More than half of households had at least seven members (52.5%). The majority were non-livestock farmers (70.4%). Approximately two-thirds had access to drinking water (65.1%), while 46.7% had unimproved latrines. One-third belonged to each socioeconomic category (Table 1).

**Table 1.** Sociodemographic characteristics of surveyed households (n=634)

| Variable | n=634, frequency (%) |
| --- | --- |
| Sex |  |
| Male | 314 (49.5) |
| Female | 320 (50.5) |
| Age group |  |
| 18-35 years | 368 (58.0) |
| 36-59 years | 221 (34.9) |
| 60+ years | 45 (7.1) |
| Education level |  |
| None | 28 (4.4) |
| Primary | 68 (10.7) |
| Secondary | 320 (50.5) |
| University | 218 (34.4) |
| Household status |  |
| Non-livestock farmer | 446 (70.4) |
| Livestock farmer | 188 (29.6) |
| Access to drinking water |  |
| Yes | 413 (65.1) |
| No | 221 (34.9) |
| Toilet type |  |
| Improved | 338 (53.3) |
| Unimproved | 296 (46.7) |

### Knowledge, Attitudes, and Practices

Only 42.9% of households had satisfactory knowledge, while 57.1% had insufficient knowledge. Although 88.5% reported having heard of zoonotic diseases, this awareness remained superficial. Mpox (78.2%) and Ebola (66.6%) were relatively well recognized, while rabies was identified by only 39.2%. Chronic diseases such as brucellosis, anthrax, or avian influenza were mentioned by only 6.6% of participants.

Attitudes were predominantly favorable (63.6%). The vast majority (95.4%) agreed that hygiene prevents zoonoses, and 85.8% recognized that animal vaccination protects families. However, 72.2% considered prayers/traditions more effective than medical prevention.

Preventive practices were very low: only 11.7% demonstrated good overall practices. While 66.6% avoided undercooked meat, only 24.9% washed hands after animal contact, 15.0% correctly disposed of carcasses, and 45.1% adopted appropriate behavior after animal bites.

**Table 2.** Knowledge, Attitude and Practice items of zoonoses among respondents (n=634)

| **Knowledge item** | Correct n(%) | Incorrect n(%) |
| --- | --- | --- |
| Ever heard of zoonotic diseases | 561 (88.5) | 73 (11.5) |
| Knows rabies as zoonosis | 248 (39.1) | 386 (60.9) |
| Knows Ebola as zoonosis | 422 (66.6) | 212 (33.4) |
| Knows Mpox as zoonosis | 496 (78.2) | 138 (21.8) |
| Knows bite/scratch transmits | 269 (42.4) | 365 (57.6) |
| Knows raw meat/milk transmits | 445 (70.2) | 189 (29.8) |
| Knows animal vaccination prevents | 417 (65.8) | 217 (34.2) |
| Knows isolating sick animals | 386 (60.9) | 248 (39.1) |
| Knows thorough cooking/boiling | 435 (68.6) | 199 (31.4) |
| Overall knowledge score | 272 (42.9) | 362 (57.1) |
| **Attitude item** |  |  |
| Zoonoses are a threat to community | 592 (93.4) | 42 (6.6) |
| My household is at real risk | 211 (33.3) | 423 (66.7) |
| Animal vaccination protects family | 544 (85.8) | 90 (14.2) |
| Hygiene prevents zoonoses | 605 (95.4) | 29 (4.6) |
| Prayers/traditions more effective* | 458 (72.2) | 176 (27.8) |
| Overall attitude score | 403 (63.6) | 231 (36.4) |
| **Practice item** |  |  |
| Avoids undercooked meat | 422 (66.6) | 212 (33.4) |
| Washes hands after animal contact | 158 (24.9) | 476 (75.1) |
| Correct disposal of carcasses | 95 (15.0) | 539 (85.0) |
| Never consumed meat from sick animal | 405 (63.9) | 229 (36.1) |
| Appropriate action after bite | 286 (45.1) | 348 (54.9) |
| Overall practice score | 74 (11.7) | 560 (88.3) |

\*Items reversed before scoring.

**Table 3.** KAP scores according to Sociodemographic characteristics (n=634)

| Variable | n | Knowledge | Statistics<br>($\chi^2/z$) | P | Attitude score | Statistics<br>($\chi^2/z$) | P | Practice<br>score | Statistics<br>($\chi^2/z$) | P | KAP score | Statistics<br>($\chi^2/z$) | P |
| --- | --- | --- | --- | --- | --- | --- | --- | --- | --- | --- | --- | --- | --- |
| Age |  |  |  |  |  |  |  |  |  |  |  |  |  |
| 18-35 years | 368 | 12 (8.5–16) | 30.77** | 0.0001 | 4 (3.625–4.25) | 6.19** | 0.045 | 2 (1–3) | 0.87** | 0.647 | 18 (14.06–22.44) | 26.81** | 0.0001 |
| 36-59 years | 221 | 11 (8–15) |  |  | 4 (3.625–4.375) |  |  | 2 (1–3) |  |  | 17.13 (13.25–21.75) |  |  |
| 60+ years | 45 | 6 (3–12) |  |  | 3.63 (3.25–4.25) |  |  | 2 (2–3) |  |  | 11 (8.25–17.75) |  |  |
| Sex |  |  |  |  |  |  |  |  |  |  |  |  |  |
| Male | 314 | 13 (10–16) | 5.42* | <0.001 | 4.13 (3.75–4.38) | 4.07* | <0.001 | 2 (1–3) | -1.57* | 0.117 | 18.31 (15.13–22.5) | 4.50* | <0.001 |
| Female | 320 | 10 (6–14) |  |  | 3.88 (3.5–4.25) |  |  | 2 (1–3) |  |  | 16.25 (11.56–20.81) |  |  |
| Education level |  |  |  |  |  |  |  |  |  |  |  |  |  |
| None | 28 | 5 (2–9.5) | 108.17** | 0.0001 | 3.5 (3.25–3.56) | 59.44** | 0.0001 | 2 (1–2) | 23.55** | 0.0001 | 10.25 (6.63–15.5) | 113.84** | 0.0001 |
| Primary | 68 | 8 (4–12.5) |  |  | 3.69 (3.25–4.13) |  |  | 2 (1–2.5) |  |  | 13 (9.19–17.88) |  |  |
| Secondary | 320 | 10 (7–14) |  |  | 4 (3.63–4.25) |  |  | 2 (1–3) |  |  | 16.25 (12.5–20.5) |  |  |
| University | 218 | 14 (11–17) |  |  | 4.13 (3.88–4.38) |  |  | 3 (2–3) |  |  | 20.44 (17–23.44) |  |  |
| Household status |  |  |  |  |  |  |  |  |  |  |  |  |  |
| Non-livestock farmer | 446 | 11 (8–15) | 3.05* | 0.002 | 4 (3.63–4.38) | 0.43* | 0.666 | 2 (1–3) | 0.41* | 0.683 | 18.56 (14.25–23) | 2.58* | 0.0098 |
| Livestock farmer | 188 | 13 (8–16) |  |  | 4 (3.63–4.31) |  |  | 2 (1–3) |  |  | 16.94 (12.88–21.25) |  |  |
| Socioeconomic status |  |  |  |  |  |  |  |  |  |  |  |  |  |
| Poor | 209 | 9 (5–14) | 55.71** | 0.0001 | 3.75 (3.25–4.25) | 41.25** | <0.001 | 2 (1–3) | 31.59** | <0.001 | 14.5 (10.5–20) | 68.10** | <0.001 |
| Medium | 209 | 12 (7–15) |  |  | 4 (3.63–4.25) |  |  | 2 (1–3) |  |  | 17.13 (12.75–21.75) |  |  |
| Rich | 208 | 13 (11–16) |  |  | 4.13 (3.88–4.44) |  |  | 3 (2–3) |  |  | 19.94 (16.88–23.25) |  |  |
| Access to drinking water |  |  |  |  |  |  |  |  |  |  |  |  |  |
| Yes | 413 | 13 (10–16) | 11.53* | <0.001 | 4.13 (3.75–4.38) | 7.29* | <0.001 | 2 (2–3) | 6.85* | <0.001 | 20.13 (16.13–23.25) | 11.99* | <0.001 |
| No | 221 | 8 (5–11) |  |  | 3.75 (3.25–4.13) |  |  | 2 (1–2) |  |  | 13.75 (10.25–16.88) |  |  |
| Toilet type |  |  |  |  |  |  |  |  |  |  |  |  |  |
| Improved | 338 | 13 (10–16) | 5.44* | <0.001 | 4.13 (3.75–4.38) | 5.08* | <0.001 | 2 (1–3) | 4.07* | <0.001 | 18.63 (15.75–22.5) | 6.25* | <0.001 |
| Unimproved | 296 | 10 (6–15) |  |  | 3.88 (3.38–4.25) |  |  | 2 (1–3) |  |  | 15.38 (10.94–20.81) |  |  |

Abbreviation: KPA=Knowledge, Attitude, and Practice. The normality test was performed for knowledge, attitude, practice scores, and the overall KAP score. As the distribution of these variables did not follow a normal distribution, the results are presented as median and interquartile range (IQR), and nonparametric tests were used for comparisons \*= Mann-Whitney test \*\* = Kruskal-Wallis test,

**Table 4.** Factors associated with satisfactory overall KAP (multivariate analysis)

| Variable | Good | Poor | OR (95% CI) | p |
| --- | --- | --- | --- | --- |
| Knowledge |  |  |  |  |
| Sex |  |  |  |  |
| Male | 159 | 155 | 1,57(1,07-2,29) | 0,020 |
| Female | 113 | 207 | Ref |  |
| Education level |  |  |  |  |
| None | 5 | 23 | Ref | - |
| Primary | 17 | 51 | 0,9 (0,27-3,21) | 0,92 |
| Secondary | 109 | 209 | 1,42 (0,46-4,39) | 0,53 |
| University | 141 | 79 | 4,54 (1,38-14,9) | 0,013 |
| Socioeconomic status |  |  |  |  |
| Poor | 63 | 146 | Ref | - |
| Medium | 90 | 119 | 0,70 (0,42–1,14) | 0,06 |
| Rich | 119 | 89 | 0,63 (0,35–1,11) | 0,045 |
| Household status |  |  |  |  |
| Non-livestock farmer | 176 | 270 | Ref | - |
| Livestock farmer | 96 | 92 | 1.94 (1,28-2,94) | 0,002 |
| Access to drinking water |  |  |  |  |
| Yes | 238 | 175 | 8,3 (5,17-13) | 0,000 |
| No | 34 | 187 | Ref | - |
| Attitude |  |  |  |  |
| Age |  |  |  |  |
| 18-35 years | 237 | 131 | Ref | - |
| 36-59 years | 147 | 74 | 1,60 (1,07-2,39) | 0,022 |
| 60+ years | 19 | 26 | 0,85 (0,40-1,76) | 0,66 |
| Sexe |  |  |  |  |
| Male | 220 | 94 | 1,57 (1,09-2,25) | 0,014 |
| Female | 183 | 137 | Ref | - |
| Education level |  |  |  |  |
| None | 4 | 24 | Ref | - |
| Primary | 31 | 37 | 4,02 (1,21-13,31) | 0,023 |
| Secondary | 199 | 119 | 7,6 (2,45-23,57) | <0,001 |
| University | 169 | 51 | 12,13 (3,74-39,32) | <0,001 |
| Access to drinking water |  |  |  |  |
| Yes | 304 | 109 | 2,85 (1,97-4,11) | <0,001 |
| No | 99 | 122 | Ref | - |
| Practice |  |  |  |  |
| Sexe |  |  |  |  |
| Male | 27 | 287 | Ref | - |
| Female | 47 | 273 | 1,92 (1,15-3,21) | 0,012 |
| Access to drinking water |  |  |  |  |
| Yes | 66 | 347 | 4.07 (1,88-8,80) | 0,000 |
| No | 8 | 213 | Ref | - |
| Type of latrine |  |  |  |  |
| Improved | 55 | 283 | 2,19 (1,24-3,87) | 0,007 |
| Unimproved | 19 | 277 | Ref | - |
| KAP Global |  |  |  |  |
| Education level |  |  |  |  |
| None | 1 | 27 | Ref | - |
| Primary | 11 | 57 | 3,58 (0,42-30,60) | 0,21 |
| Secondary | 70 | 248 | 4,33 (0,55-33,83) | 0,16 |
| University | 106 | 114 | 12,06 (1,54-94,18) | 0,018 |
| Access to drinking water |  |  |  |  |
| Yes | 171 | 242 | 6,74(3,91-11,60) | <0,001 |
| No | 17 | 204 | Ref | - |

Abbreviation: OR=odds ratio; CI=confidence interval; KAP=Knowledge, Attitude, and Practice.

## Discussion

The findings of this study demonstrate that households in the urban and peri-urban health zones of Bukavu exhibit limited overall knowledge, generally positive attitudes, but remarkably poor preventive practices regarding priority zoonotic diseases, highlighting significant vulnerability to zoonotic health threats in this setting. The overall level of satisfactory knowledge regarding zoonoses remained low among surveyed households (42.9%). Although a large majority of participants (88.5%) reported having heard of zoonotic diseases, this awareness was predominantly superficial and highly selective. Highly publicized emerging viral diseases, such as Mpox (78.2%) and Ebola (66.7%), were widely recognized. In contrast, endemic zoonoses such as rabies were identified by less than half of the respondents (39.2%), while chronic or neglected zoonotic conditions (including brucellosis, anthrax, and avian influenza) were rarely mentioned (6.6%).

In the context of Bukavu, this disparity in knowledge can be explained by the nature of public health communication and recent epidemiological events. South Kivu province has experienced recurring major health emergencies, including the recent 2023–2024 Mpox outbreak(42) and threat of Ebola transmission (43) , which triggered intensive media coverage, community sensitization campaigns, and international humanitarian responses(44). Conversely, endemic diseases like rabies or brucellosis receive negligible routine health promotion, leading to reactive rather than comprehensive community knowledge.(41) Consequently, public understanding is heavily shaped by crisis-driven communications rather than structured, continuous health education.

These observations align with studies conducted in other sub-Saharan African contexts. In South Africa, Ngoshe et al. reported adequate knowledge in 53.2% of communal livestock farmers but highlighted key gaps in non-epidemic zoonoses.(45) Similarly, selective knowledge was observed in Ethiopia by Alemayehu et al. (54.2%) (35) and in Cameroon by Zangue et al., who documented very low overall knowledge scores among farmers(37) . Across these settings, disease familiarity remains heavily dependent on public visibility and recent outbreak history.(46)

Multivariable analysis revealed that education level was a primary determinant of knowledge, with university-educated respondents being significantly more likely to possess satisfactory knowledge (OR = 4.54; 95% CI: 1.38–14.9). Higher formal education enhances health literacy, information comprehension, and critical evaluation of disease transmission risks (47). Furthermore, livestock-owning households demonstrated higher knowledge scores than non-livestock owners (OR = 1.94; 95% CI: 1.28–2.94), likely due to direct occupational exposure and increased awareness of animal health issues (35).

Beyond disease awareness, household attitudes toward zoonotic disease prevention were generally favorable (63.6%), though highly nuanced. Most respondents agreed that general hygiene prevents zoonotic transmission (95.4%) and that animal vaccination protects household health (85.8%). However, a substantial proportion (72.2%) considered traditional or religious practices more effective than modern medical prevention. This duality reflects the influence of socio-cultural norms in Bukavu, where community perception of illness often combines scientific understanding with traditional beliefs. Similar favorable attitudes coupled with persistent socio-cultural beliefs have been reported in Ethiopia by Abunna et al. and Alemayehu et al., who noted that declared adherence to preventive principles is frequently driven by social desirability rather than thorough biological understanding(35,48). Access to drinking water was also positively associated with favorable attitudes (OR = 2.85; 95% CI: 1.97–4.11), suggesting that better environmental infrastructure fosters greater perceived self-efficacy in disease prevention(49).

Despite favorable attitudes and moderate knowledge, safe preventive practices were alarmingly low in Bukavu, with only 11.7% of households demonstrating adequate protective behaviors. While basic culinary hygiene measures such as avoiding undercooked meat were relatively common (66.6%), critical protective practices remained poorly adopted: only 24.9% of participants reported washing hands with soap after animal contact, 15.0% disposed of animal carcasses correctly, and 45.1% sought appropriate healthcare following animal exposure or bites. This pronounced gap between positive attitudes and actual preventive practices is a recurring finding in KAP literature on zoonotic diseases in low-resource settings(35,45). In Bukavu, this discrepancy is driven by structural and economic determinants rather than individual negligence. Even when individuals understand transmission risks, acute water scarcity and inadequate sanitation facilities make routine handwashing and hygienic waste disposal practically unfeasible (50,51). Furthermore, economic hardship may compel households to salvage meat from dead or sick animals or forego veterinary services due to cost (35,37).

Multivariable logistic regression confirmed that structural factors were the strongest predictors of good preventive practices. Households with access to drinking water were over four times more likely to adopt adequate practices (OR = 4.07; 95% CI: 1.88–8.80), and those with improved sanitation latrines showed higher adoption rates (OR = 2.19; 95% CI: 1.24– 3.87). Female gender was also positively associated with adequate practices (OR = 1.92; 95% CI: 1.15–3.21), underscoring women’s primary role in domestic hygiene, food preparation, and household caregiving in this setting(37,52). Overall, these findings highlight that health education campaigns alone cannot bridge the practice gap without parallel investments in WASH infrastructure and structural poverty reduction within a One Health framework.(40,52,53)

Collectively, these findings demonstrate that health education campaigns alone cannot bridge the practice gap without parallel structural investments(53). Addressing zoonotic risks in dense urban settings like Bukavu requires an operationalized One Health strategy that integrates human public health, veterinary care, and urban environmental infrastructure:

1. WASH and Environmental Infrastructure: Priorities must focus on expanding urban drinking water access and waste management systems, as structural WASH availability is a prerequisite for preventive behavior compliance.
2. Community-Based Veterinary Services: Affordable rabies vaccination campaigns for domestic dogs and accessible veterinary extension services are needed to reduce zoonotic spillover at the animal source.
3. Targeted Risk Communication: Health education should move beyond emergency epidemic response (Ebola/Mpox) to address neglected endemic zoonoses (rabies, brucellosis) while integrating local socio-cultural beliefs into risk communication frameworks.

In the context of the dense urban environment of Bukavu, this study presents several methodological and structural limitations that should be considered when interpreting the findings. First, the cross-sectional design restricts the ability to establish temporal or causal relationships between sociodemographic or environmental determinants and household KAP levels. Second, self-reported data regarding health behaviors and preventive measures are susceptible to social desirability and recall biases, which may lead to an overestimation of favorable practices. Third, security instability and geographic constraints in certain sub-quarters of Bagira and Kadutu restricted access during field collection, potentially introducing minor sampling bias. Finally, while the study evaluated major priority zoonoses in Eastern DRC, biological sampling of household domestic animals was not conducted, precluding direct correlation between reported KAP scores and objective pathogen prevalence.

In conclusion, this study demonstrates a profound structural mismatch between disease awareness, attitudes, and actual preventive behaviors regarding priority zoonoses among Bukavu households. Although public health communication during recent Ebola and Mpox epidemics has successfully heightened general disease visibility, low compliance with safe protective behaviors such as post-exposure healthcare seeking and hygienic carcass management leaves urban communities highly vulnerable to zoonotic spillover. This gap is primarily driven by systemic environmental and socioeconomic barriers, particularly the lack of accessible drinking water and adequate sanitation infrastructure, rather than a mere lack of health literacy. Addressing these challenges requires transitioning from isolated emergency risk communication toward an integrated One Health framework. Priority intervention strategies must combine targeted health education on neglected endemic zoonoses, such as rabies and brucellosis, with structural investments in urban WASH infrastructure and accessible community-based veterinary services.

## Data Availability

Data from this study are available upon reasonable request.

## Declarations

### Ethics Approval and Consent to Participate

The study was conducted according to the Declaration of Helsinki. Informed consent was obtained from all participants. Confidentiality and anonymity were guaranteed.

